# Novel insights into the common heritable liability to addiction: a multivariate genome-wide association study

**DOI:** 10.1101/2021.11.19.21266548

**Authors:** Tabea Schoeler, Jessie Baldwin, Andrea Allegrini, Wikus Barkhuizen, Andrew McQuillin, Nicola Pirastu, Zoltán Kutalik, Jean-Baptiste Pingault

## Abstract

Addiction to nicotine, alcohol and cannabis commonly co-occurs, which is thought to partly stem from a common heritable liability. To elucidate its genetic architecture, we modelled the common liability to addiction, inferred from genetic correlations among six measures of dependence and frequency of use of nicotine, alcohol and cannabis. Forty-two genetic variants were identified in the multivariate genome-wide association study on the common liability to addiction, of which 67% were novel and not associated with the six phenotypes. Mapped genes highlighted the role of dopamine (e.g., dopamine D2 gene), and showed enrichment for several components of the central nervous systems (e.g., mesocorticolimbic brain regions) and molecular pathways (dopaminergic, glutamatergic, GABAergic) that are thought to modulate drug reinforcement. Genetic correlations with other traits were most prominent for reward-related behaviours (e.g., risk-taking, cocaine and heroin use) and mood (e.g., depression, insomnia). These genome-wide results triangulate and expand previous preclinical and human studies focusing on the neurobiological substrates of addiction, and help to elucidate the common genetic architecture underlying addiction.

## Introduction

Addiction is one of the leading causes of preventable harms^1^, and considerable research efforts are made to better understand its aetiology. Addiction is typically not restricted to just one class of substance, as dependence of accessible psychoactive substances such as nicotine, alcohol and cannabis often co-occurs^2^. This co-occurring pattern of use has been shown to be particularly detrimental to the individual and to society as a whole^3^.

Aetiological models posit that addiction to multiple substances stems from a common liability to addiction^4,5^ – a latent continuous trait accounting for the shared risk of developing addiction to different substances. Based on findings from genomic^6–8^ and behavioural genetic studies^9,10^, it is assumed that this common liability includes a genetic component. Indeed, genetic correlations between use of different classes of psychoactive substances are substantial, as estimated in twin (up to *r*_g_∼0.89^11–13^) and genome-wide association (GWA) studies (up to *r*_g_∼0.70^7,14–16^). The underlying molecular mechanisms of this common heritable liability to addiction are, however, not fully understood. While it has been shown that genotypic variations contribute to the common heritable liability to addiction^17^, an investigation into the specific genome-wide effects has yet to be conducted. Furthermore, although increasingly large GWA studies have identified growing numbers of genetic risk variants associated with individual substance use phenotypes^7^, it remains unclear as to whether associated risk variants reflect shared (common) or non-shared (substance specific) risk across different addiction phenotypes. Some identified genetic variants likely operate through substance-specific pharmacological pathways, as is the case for variants affecting nicotinic receptors (e.g. genes coding for nicotinic acetylcholine receptors, such as *CHRNA3*-*CHRNA5*^18^) or alcohol metabolism (e.g. variants in the alcohol dehydrogenases gene family, such as *ADH1B*^7,15^, *ADH1C*^14,15^). Other variants may affect common pathways, such as variants associating with two or more classes of psychoactive substances (e.g. *BDNF*^7,19,20^, *PDE4B*^7,20,21^ or *DRD2*^7,15,18,20,22^) or behavioural phenotypes (e.g., the top variant identified for cannabis use disorder, which also associates with ADHD and risk-taking^23^).

The aforementioned genetic overlap complicates research on causes and consequences of addiction, and distilling shared (common) from non-shared (substance specific) genetic risk is pivotal to the interpretation of genome-wide discoveries. One way of scrutinizing putative pleiotropic variants is to explicitly model the genetic overlap among different phenotypes indexing addiction, using multivariate methods such as genomic structural equation modelling (genomic SEM^24^). Applying this method has already been helpful in characterising shared genetic influences across dimensions of psychopathology^24–31^ and cognition^32–35^. In addition to assessing shared effects of suspected pleiotropic variants, genomic SEM also has the potential to identify novel genetic variants not previously identified in univariate GWA studies on individual phenotypes^29^. This is expected, since shared risk is thought to be expressed indirectly via the common liability, resulting in inherently small effects, which hampers detection of pleiotropic variants in univariate GWA analyses. A multivariate GWA can therefore boost discovery of shared variants directly associated with a common heritable liability. While genetically informed methods using polygenic scores have already explored risk factors involved in the common liability to addiction^36,37^, a multivariate GWA of the shared and non-shared genetic architecture can further deepen our understanding of biological pathways underlying addiction to multiple substances. Indeed, leveraging genetically informed methods would allow us to revisit long-theorized biological pathways underlying addiction, and to triangulate evidence from behavioural genetic^11^, brain imaging^38^, candidate gene^39^ and preclinical studies^40^ focusing on the role of genetics and neural substrates of addiction. Together, such triangulated findings would help researchers and clinicians to better understand biological and developmental pathways involved in risk of developing addiction to commonly used and abused psychoactive substances.

To unravel the genetic architecture underlying addiction to nicotine, alcohol and cannabis, here we conduct a multivariate GWA analysis on their common heritable liability. To model the common liability to addiction to these substances, we include phenotypes indexing clinical (diagnosis of dependence) as well as quantitative (frequency of use) measures of addiction to nicotine, alcohol and cannabis. More specifically, we conduct a multivariate GWA of the common heritable liability, with the aim to

a. identify putative genetic variants associated with the common liability (i.e., shared/pleiotropic variants) and variants specific to the use of different classes of substances (i.e., non-shared)
b. characterize the functional features of genetic variants associated with the common liability
c. assess the genetic correlations between the common liability with other complex traits
d. evaluate the validity of the causal claims imposed by a common liability model of addiction

## Materials and methods

### GWA summary datasets

We screened GWA summary statistics of addiction-related phenotypes for the most commonly used and misused psychoactive substances, namely nicotine, alcohol and cannabis. For each substance class, we included one clinical (diagnosis of dependence) and one quantitative (frequency of use) measure of addiction. The following summary statistics were included, derived from samples of individuals of European ancestry: Data on alcohol use disorder (n=28,757)^14^ and cannabis use disorder (n=358,534)^23^ was obtained from the Substance Use Disorders working group of the Psychiatric Genomics Consortium (PGC-SUD). Nicotine dependence (n=244,890) was taken from GWAS ATLAS^41^ (cf. URLs). Frequency of cigarette (n=245,876) and alcohol use (n=513,208), obtained from the GWA meta-analysis by Liu et al.^7^. The phenotypes were measured as the number of cigarettes/alcoholic drinks consumed during periods of consumption (assessed through the questions “How many cigarettes do/did you smoke per day?” and “How many drinks do/did you have each week/month?”). Frequency of cannabis use (n=24,798, from the question “Considering when you were taking cannabis most regularly, how often did you take it?”) was obtained from the Neale Lab UKBB summary statistics (cf. **URLs**). Additional details of each of the included summary statistic files can be found in **sTable 1** (Supplement). As the included phenotypes are considered to reflect different dimensions of addiction, we coined their shared genetic architecture the ‘common heritable liability to addiction’.

### Genomic model of the common heritable liability to addiction

We first estimated the genetic correlations (*r*_*g*_) among the individual phenotypes using genomic structural equation modelling (genomic SEM^24^) version 0.0.3. The method uses an extension of LD-score regression^42^ and accounts for sample overlap across studies through the LD-score intercept. In confirmatory factor analysis, we fitted a structural equation model with a single latent factor, representing a common liability to addiction to nicotine, alcohol and cannabis, onto which the six indicators loaded **(Figure 1b)**. Equality constrains were imposed on paths belonging to the same pattern of substance use, i.e., equal weights across measures of dependence, and equal weights across measures of frequency of use. Correlated residuals were included to allow for within-substance class associations, as depicted in **Figure 1b**. The Diagonally Weighted Least Squares (DWLS) estimator was used and model fit was assessed based on the Comparative Fit Index (CFI) and the standardized root mean square residual (SRMR).

**Figure 1.**
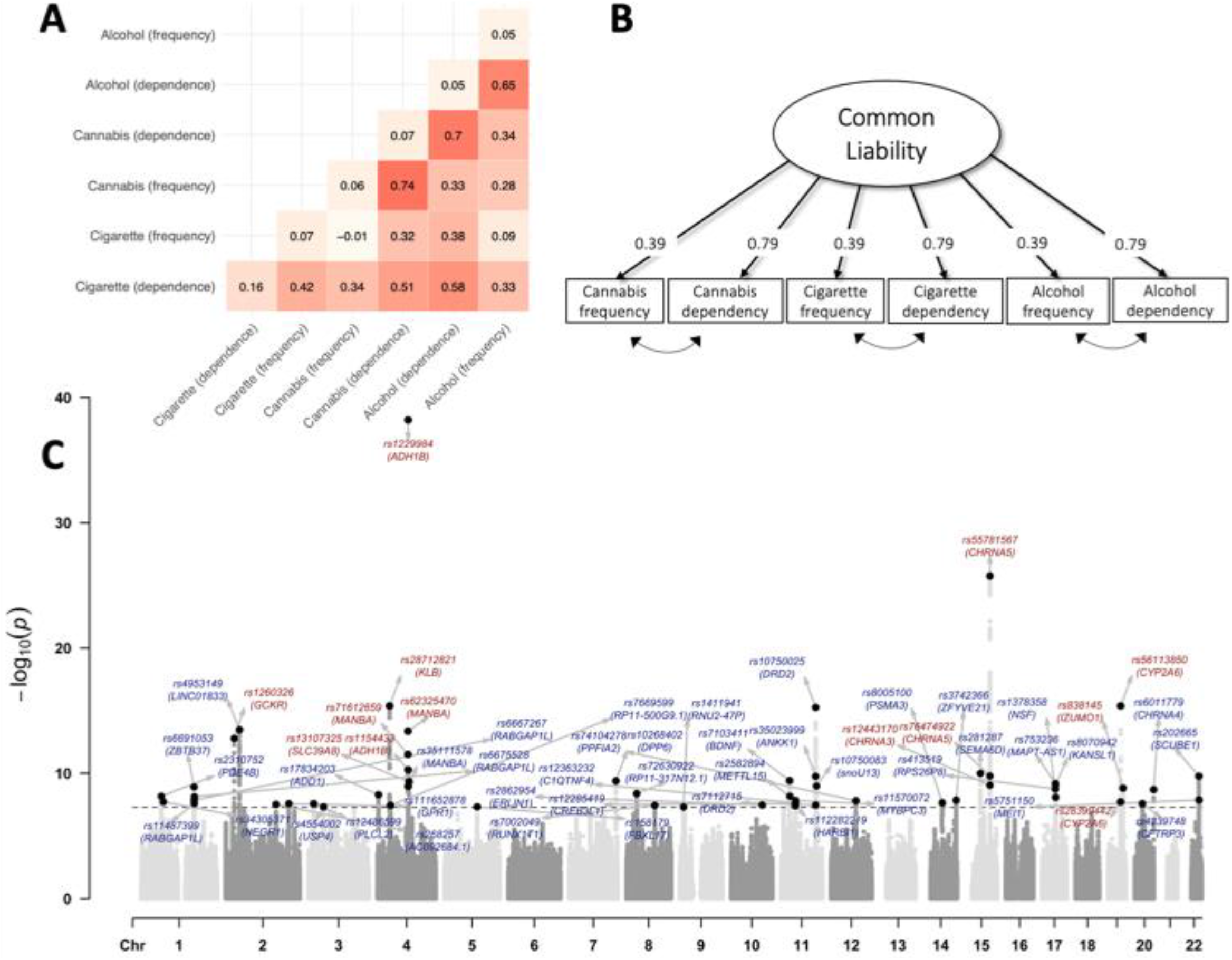
Multivariate genome-wide association study of the shared genetic architecture of cigarette, alcohol and cannabis use **Panel A**. Heat map displaying the genetic correlations among the six substance use phenotypes. Shown are the genetic correlations between each of the cigarette, alcohol and cannabis phenotypes, with SNP-heritability estimates displayed down the diagonal. The mean genetic correlation is *r*_*g*_=0.4 [sd=0.21, median=0.34 and range (−0.01-0.74)]. **Panel B**. Genomic structural equation model fitted on the genetic covariation matrices of the individual cigarette, alcohol and cannabis use phenotypes. Squares represent observed variables (the measured cigarette, alcohol and cannabis use phenotypes). The circle represents the latent variable, i.e., the common heritable liability to addiction, derived through factor analysis of the genetic correlations between the individual substance use phenotypes. Single-headed arrows are regression paths constrained to be equal across measures of frequency of use and dependence. **Panel C**. Manhattan plot of the SNP effects obtained from the multivariate genome-wide association analysis on the common liability. Labels are provided for the LD-independent genome-wide significant SNPs (i.e., SNPs above the horizontal line, with *p*<5×10^−8^) and gene names obtained through positional mapping. The x-axis refers to chromosomal position, the y-axis refers to the *p*-value on a -log10 scale. Genetic variants coloured in red index variants that showed heterogeneous effects across the individual cigarette, alcohol and cannabis use phenotypes (Q_SNP_ *p*<5×10^−8^), indicating that their effects operate not entirely through the common liability. Genetic variants coloured in blue index genetic variants that did not show heterogeneous effects across the individual cigarette, alcohol and cannabis use phenotypes (Q_SNP_ *p*≥5×10^−8^), indicating that their effects are likely to operate through the common liability.

### Multivariate genome-wide association analysis

For the multivariate GWA on the common heritable liability to addiction, the summary statistics for the individual addiction phenotypes were restricted to single nucleotide polymorphisms (SNPs) contained in the 1000 genomes phase 3 reference sample (with a minor allele frequency (MAF) > 1%) and SNPs that were present in all GWA summary datasets included in the analysis. Genomic control was applied to all summary statistics showing evidence of uncontrolled confounding (LD score intercept > 1), by multiplying standard errors by the LD score intercept. To identify lead SNPs after conducting the GWA on the common liability, we selected LD-independent SNPs (*r*^2^<0.1 within 250 kb) based on genome-wide significance (*p*<5×10^−8^).

To determine whether the effects of the identified lead SNPs are likely to act through the common liability, we applied the heterogeneity test as implemented in Genomic SEM. The resulting Q-statistic (Q_SNP_) is a *χ*^*2*^distributed test statistic, with significant Q_SNP_ estimates (*p*<5×10^−8^) indicating that the SNP effect does not act entirely through the common liability. Based on Q_SNP_, we selected only SNPs that did not show evidence of heterogeneity (Q_SNP_ *p*≥5×10^−8^) before conducting functional follow-up analyses of SNPs associated with the common liability.

Two complementary strategies were used to map SNPs to genes: (i) positional mapping of lead SNPs and (ii) expression quantitative trait loci (eQTL) mapping of lead SNPs. For positional mapping (i), g:ProfileR^43,44^ and Phenoscanner^45^ were used, mapping SNPs to genes based on being physically located inside a gene. SNPs identified in GWA studies often reside in non-coding regions of the genome (intergenic and intronic), but can nevertheless influence gene expression of nearby genes. We therefore applied (ii) eQTL mapping on the lead SNPs (*cis*-eQTL, variants in a +/- 1 megabase window around the transcription start site of a given gene) using Qtlizer^46^. Qtlizer integrates a number of eQTL databases (e.g. GTEx Portal^47^, Haploreg^48^, GRASP^49^, GEUVADIS^50^, SCAN^51^, seeQTL^52^, Blood eQTL Browser^53^, pGWAS^54^, ExSNP^55^ and BRAINEAC^56^), to assign noncoding SNPs to their cognate genes.

Finally, to explore previously identified associations of lead SNPs with other phenotypes, we searched the PhenoScanner^45^, a database of genotype-phenotype associations from existing GWA studies. For comparability, the same set of analyses was employed to characterise SNPs associated with the individual addiction phenotypes, including only lead SNPs that did not solely operate through the common liability (Q_SNP_ *p*<5×10^−8^). That way, the results obtained from gene annotation and functional analyses are interpretable with respect to shared (i.e., effects operating through the common liability) and non-shared (i.e., effects specific to the individual addiction phenotypes) risks.

### Pathway enrichment analysis of genetic variants associated with the common liability

To identify the most likely biological pathways underlying the common heritable liability to addiction, we used Data-driven Expression-Prioritized Integration for Complex Traits (DEPICT^57^) and Pathway SCoring ALgorithm (PASCAL)^58^. DEPCIT was used to test for tissue/cell type enrichment of a set of LD-independent SNPs (*r*^2^<0.05 within 500 kb) outside genome-wide significance (*p*<5×10^−5^). PASCAL was used to test for enrichment of all SNPs, using three gene sets (BIOCARTA, KEGG, REACTOME) curated by the Molecular Signatures Database (MSigDB^59^) and gene sets defined by DEPICT. Prior to running the analyses, the GWA on the common liability was filtered according to the Q_SNP_ statistic, retaining only SNPs operating through the common liability (Q_SNP_ *p*≥5×10^−8^). Results were corrected for multiple testing using false discovery rate (FDR) correction (controlled at 5%). Further details regarding the application of the two methods can be found in the Supplement (sMethods).

### Genetic correlations between the common liability and other complex traits

Bivariate LD score regression analyses were performed in Genomic SEM, to estimate the genetic correlations between SNPs operating through the common liability (i.e., SNPs with Q_SNP_ *p*≥5×10^−8^) and 35 other traits related to physical features (e.g., height, body mass index), personality (e.g., risk-taking, neuroticism), social variables (e.g., socioeconomic status, education) and mental health (e.g., schizophrenia, depression). A complete list of the included GWA summary statistics can be found in **sTable 1** (Supplement). FDR correction (controlled at 5%) was used to adjust for multiple testing.

### Evaluation of the causal claims implied by the common liability theory

It is important to note that the common liability to addiction, constructed from the observed covariance structure of phenotypes reflecting addictive behaviours, constitutes only a statistical model. As such, the model’s fit statistics do not examine the validity of the causal claims imposed by the model. For example, due to statistical equivalence, models with distinct interpretations (e.g. network models versus latent factor models) can have equivalent fit when constructed from the same data^60^. Therefore, model fit reveals little about the underlying causal relationships between the observed and latent variables. To assess the factor model-implied causal pathways, where the common liability factor is thought to causally influence all its indicators, Mendelian Randomization (MR) analysis was used to assess the effects between the common liability and the individual substance use phenotypes. Inverse variance weighted (IVW) MR implemented in TwoSampleMR^61^ package was applied to all analyses. The genetic markers instrumenting the common liability were selected based on genome-wide significance (*p*<5×10^−8^) and Q_snp_, retaining only SNPs that operated through the common liability (Q_snp_ *p*≥5×10^−8^). To facilitate comparability of the MR estimates, the beta estimates for the included SNPs were standardized by dividing the z-scores by the square root of the sample size before conducting MR (cf. Supplement).

## Results

### Genomic model of the common heritable liability to addiction

The correlations among the individual cigarette, alcohol and cannabis use phenotypes are presented in the heatmap in **Figure 1a** (cf. Supplement, **sTable 2** for estimates). Genetic correlations varied widely between the individual substance phenotypes, ranging from *r*_g_=-0.01 to *r*_g_=0.74 (mean *r*_g_=0.40; SD=0.21). All standardized factor loadings are presented in **Figure 1b** (cf. also **sTable 3**, Supplement), showing that the constrained loadings were estimated to be 0.39 for frequency measures of substance use and 0.79 for substance dependence measures. On average, the common factor accounted for 38.81% (range 15.21%-62.41%) of the genetic variance in the six substance use phenotypes. The genomic common liability model showed evidence of a good model fit (CFI=0.97, SRMR=0.07, cf. **sTable 4**).

### Genetic variants associated with the common heritable liability

6,500,152 SNPs were included in the GWA of the common liability. Since our SNP estimates were derived from overlapping samples, we used the formula developed by Mallard et al.^62^ to derive the effective samples size (cf. sMethods, Supplement), which was estimated to be N=187,062. The Manhattan and Q-Q (quantile-quantile) plot of the common liability GWA are shown in **Figure 1c** and **sFigure 1** (Supplement), respectively. The main results for all GWA analyses, including the multivariate analysis on the common liability and the six univariate analyses on the individual substance use phenotypes are summarised in **sTable 5-7** and **sFigure 1** (Supplement). In brief, the GWA on the common liability identified 3,509 genome-wide (*p*<5×10^−8^) SNPs, tagging 55 LD-independent SNPs. After removing SNPs showing significant heterogeneity (Q_SNP_ *p*<5×10^−8^), 42 SNPs operating through the common liability remained (cf. SNPs highlighted in blue in the Manhattan plot, **Figure 1c**). Of the 42 SNPs, 28 (66.67%) were novel, i.e., have not been associated with any of the individual substance use phenotypes. Positional mapping showed that the top five SNPs (rs10750025, rs4953149, rs281287, rs202665, rs35023999) operating through the common liability lay mostly outside coding regions, located close to *DRD2, LINC01833, SEMA6D, SCUBE1* and *ANKK1*, respectively. Further inspection through eQTL mapping indicated that the aforementioned SNPs acted as eQTLs for positionally mapped genes, highlighting their putative role in the common liability via gene expression (cf. **sTable 8**, Supplement). A search in the PhenoScanner database^45^ indicated that the five lead SNPs operating through the common liability have previously been linked to a number of behavioural phenotypes, such as neuroticism, irritability, smoking status or time spent in front of the computer (cf. **sTable 9**, Supplement). Of note, 13 of the 55 SNPs associated with the common liability still showed heterogeneous effects across the individual substance use phenotypes (Q_SNP_ *p*<5×10^−8^, highlighted in red in the Manhattan plot, Figure 1c). Those SNPs can be considered as false discoveries, which may result from a single or a subset of SNPs with large effects on the individual substance use phenotypes^35^. Among all 6,500,152 SNPs included in the common liability GWA, 2,356 (0.04%) showed heterogeneous effects.

For comparison, we also evaluated the GWA results of the individual substance use phenotypes, focusing on significant variants (*p*<5×10^−8^) from the original GWA studies that showed heterogenous effects (Q_SNP_ *p*<5×10^−8^). Here, a number of variants appeared to be specific with respect to the class of substance. For alcohol use, the most prominent SNP was rs1229984, a variant on the alcohol dehydrogenase 1B gene (*ADH1B*). As shown in **Figure 2**, this variant was associated with all alcohol use phenotypes, but none of the cigarette or cannabis use phenotypes. Other identified SNPs related to the alcohol dehydrogenase group included rs1154433 and rs283412 (intron variants located on *ADH1B* and *ADH1C*, respectively) and rs1154433 acting as a *cis*-eQTL for *ADH1A* and *ADH4*. For cigarette use, a number of SNPs related to nicotinic pathways showed genome-wide effects, mapping onto nicotinic receptor genes (e.g., rs76474922 and rs58379124, intron variants located on *CHRNA5 and CHRNB3, respectively*) and variants affecting gene expression of *CHRNA2, CHRNA3, CHRNA4, CHRNA5, CHRNB4* and *CHRNB5 (*cf. *cis*-eQTLs rs8034191, rs7174367, rs76474922, rs12595350, rs2273500, rs1052035, rs72740960, rs73229090). Only two SNPs were associated with cannabis use phenotypes, of which one variant (rs7783012, FOXP2) appeared to operate via the common liability (Q_SNP_ *p*≥5×10^−8^). This is supported by studies implicating rs7783012 in a number of phenotypes related to externalising behaviours (e.g., ADHD, risk-taking and risky sexual behaviour^45^), rather than pathways pointing to the endocannabinoid system specifically.

**Figure 2.**
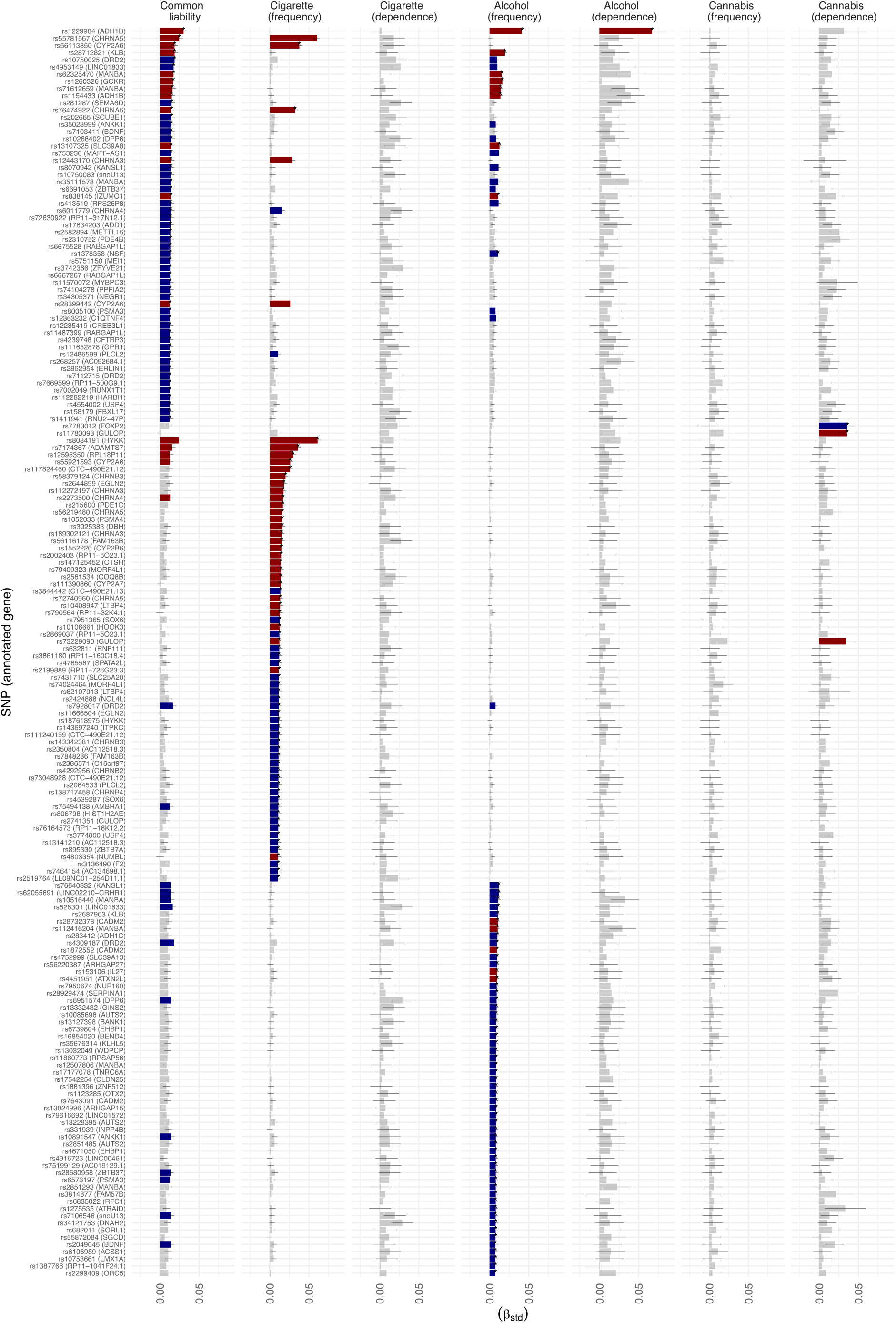
Associations of genetic variants with the common liability (blue) and the individual substance use phenotypes (red) Plotted are the standardized beta coefficients (β_std_) and their confidence intervals (cf. Supplement for details and corresponding formula) obtained from the multivariate genome-wide association (GWA) analysis on the common liability (column 1) and the univariate GWA analyses on the individual substance use phenotypes (columns 2-7). Displayed are genetic variants associated (*p*<5×10^−8^) with at least one of the individual substance use phenotypes and/or the common liability. Bars coloured in grey index genetic variants that are not significantly associated (*p*>5×10^−8^) with their respective phenotype. Bars coloured in red index genetic variants that showed heterogeneous effects across the individual cigarette, alcohol and cannabis use phenotypes (Q_SNP_ *p*<5×10^−8^), indicating that their effects operate not entirely through the common liability. Bars coloured in blue index genetic variants that did not show heterogeneous effects across the individual cigarette, alcohol and cannabis use phenotypes (Q_SNP_ *p*>5×10^−8^), indicating that their effects are unlikely to entirely operate through the common liability. The complete set of estimates can be found in sTable 7. The asterisks (*) highlight genetic variants that were identified as LD-independent SNPs following clumping

### Pathway enrichment analyses of genes associated with the common heritable liability

Testing for tissue and cell type enrichment in DEPICT revealed 22 pathways associated (with FDR controlled at 5%) with the common liability **(Figure 3A)**, which were all part of central nervous system tissues. A pattern of regional enrichment highlighted the role of a number of brain structures involved in the mesocorticolimbic brain circuits theorized to underlie addictive behaviours; notably, enrichment was present for regions involved in reward and emotion processing (limbic structures), motivation (basal ganglia), memory (hippocampus, parahippocampal gyrus, entorhinal cortex) and cognitive control (frontal lobe areas). In addition, the findings suggested that other brain structures may play a role in risk of addiction, such as areas involved in stress response (hypothalamus), or those relevant for visual processing (e.g., parahippocampal cortex, visual cortex, occipital lobe). Given this widespread network of brain areas, our results feed into theories suggesting that genetic risk to addiction is not solely the manifestation of altered limbic reward processes^63^. Since the highlighted pathways were most prominently enriched for the common liability, and less so for individual substance use phenotypes, the aforementioned brain circuits may tap into somewhat distinct features characterizing the common liability.

**Figure 3.**
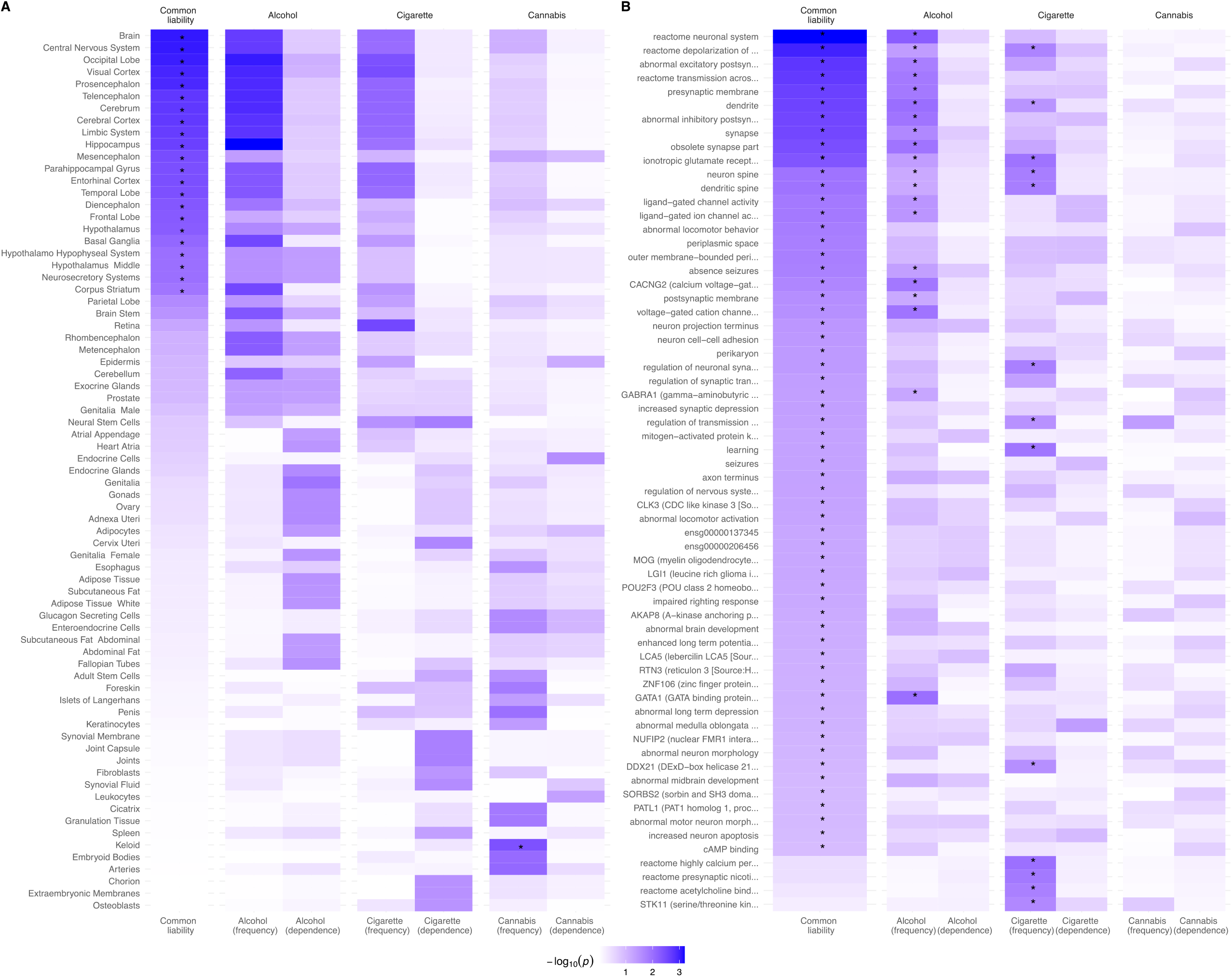
Pathway enrichment analyses of genes associated with the common heritable liability Shown are the results obtained from pathway enrichment analysis conducted in DEPICT and PASCAL. The common liability GWA results (filtered according to Q_SNP_ *p*<5×10^−8^) and the individual substance use GWA summary statistics were used as the input. The violet shading indexes the significance level corresponding to each tested pathway. The asterisk marks pathways that remained significant after correction for multiple testing (False Discovery Rate controlled at 5%). **Panel A** highlights results obtained from the tissue/cell type enrichment analysis done in DEPICT. Displayed are the -log10(*p*-value) for all pathways that were significant (*p*<0.05) in at least one of the included GWA studies. **Panel B** depicts results obtained from pathway analysis done in PASCAL, using gene-sets curated by the Molecular Signatures Database (n=1077 sets) and DEPICT (n=14462 sets). For the common liability, n=478 pathways were significant after FDR correction for multiple testing. Displayed in the figure are the 15 most significant pathways per GWA study. The full set of results is listed in **sTable 10-11** (Supplement).

Findings from PASCAL **(Figure 3B)** complement such conclusions, highlighting the role of neuronal signalling pathways in the common liability. Overall, 481 pathways were significantly (FDR controlled at 5%) enriched for the common liability, of which the top pathways related to broader categories of neurotransmitter functioning (e.g., neural system, abnormal excitatory postsynaptic currents, abnormal excitatory postsynaptic currents, transmission across chemical synapses). While the GWA analysis on the common liability identified the dopamine receptor D2 gene as the lead gene, pathway enrichment analyses implicated not only dopamine (e.g., dopamine neurotransmitter release cycle, dopamine binding), but also a number of other neurotransmitter systems, notably glutamate (e.g., ionotropic glutamate receptor complex, glutamate receptor activity), GABA (GABA-A receptor activity) or serotonin (serotonin neurotransmitter release cycle). Other identified pathways echoed the function of brain regions described above, such as those involved in learning and cognition (e.g., visual and associative learning, memory, cognition) or stress response (e.g., cAMP binding). Finally, a number of pathways were uniquely associated with the individual substance use phenotypes, mainly including those related to nicotinic pathways for cigarette use (e.g., presynaptic nicotinic acetylcholine receptors, postsynaptic nicotinic acetylcholine receptors, acetylcholine binding and downstream events). For alcohol, the ethanol oxidation pathway did not remain significant after correction for multiple testing, and there was no significant enrichment for cannabis use phenotypes.

All estimates obtained from DEPCIT and PASCAL are included in **sTable 11-12** (Supplement).

### Genetic correlations between the common liability and other complex traits

Using the input from the Q_SNP_-filtered GWA of the common liability and the GWA summary statistics for 41 traits (cf. **sTable 1** in Supplement for details), we found significant correlations with 36 complex traits after correction for multiple testing **(Figure 4)**. As expected, the largest positive correlations were present between the common liability and its “constituents”, i.e., the cigarette, alcohol and cannabis use phenotypes used to derive the common liability (mean *r*_*g*_=0.68). Among the other traits, the largest genetic correlations were present for cocaine and opioid dependence (both *r*_*g*_=0.60), number of sexual partners (*r*_*g*_=0.49), ADHD (*r*_*g*_=0.48) and risk tolerance (*r*_*g*_=0.41). Moderate genetic correlations were also present for a number of traits relating to mood, including insomnia (*r*_*g*_=0.35) and depression (*r*_*g*_=0.34). No significant (FDR controlled at 5%) associations were found with birth weight, openness, obsessive-compulsive disorder, anorexia and cortical surface area.

**Figure 4.**
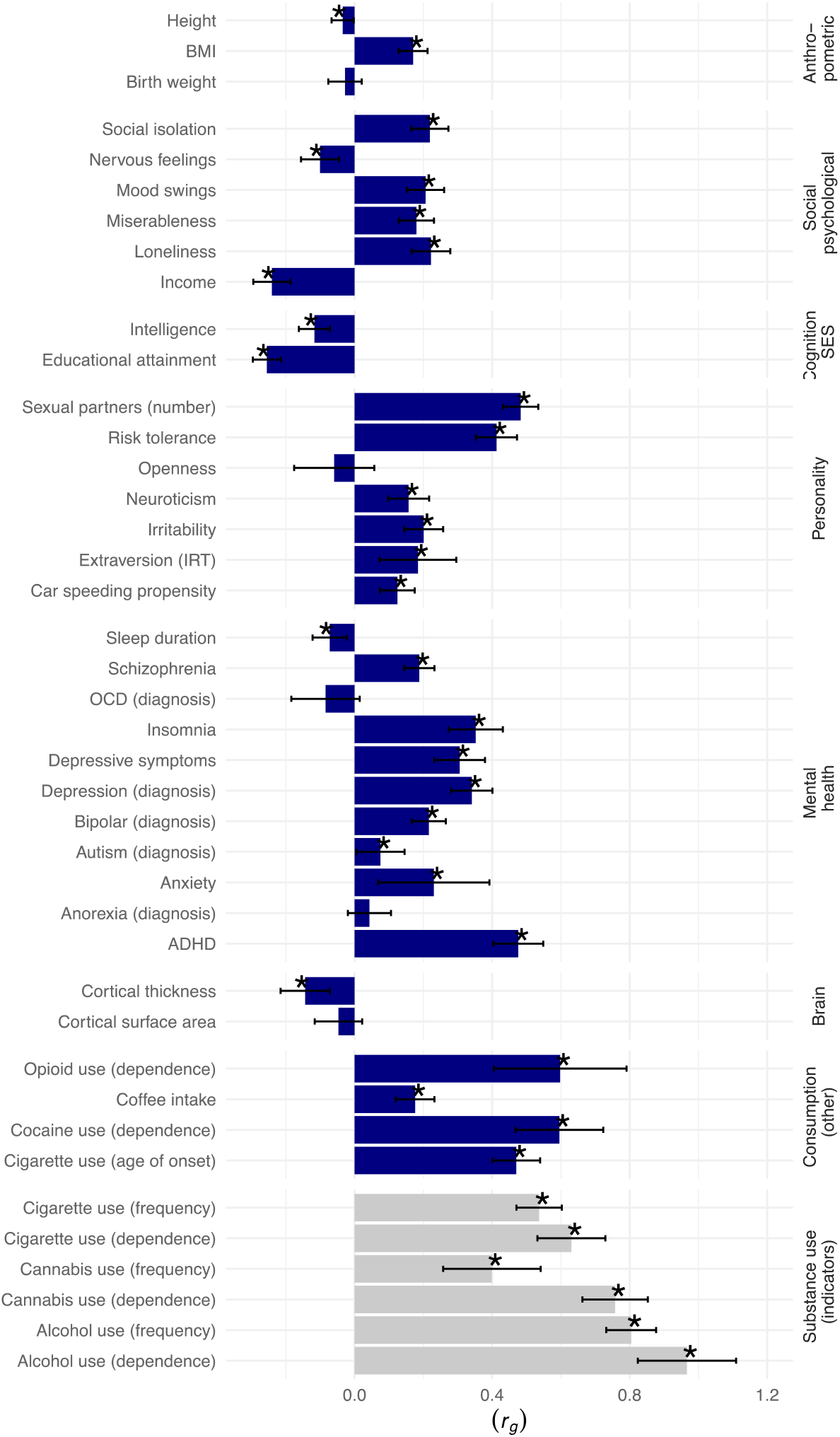
Genetic correlations between the common liability and other traits Shown are the genetic correlations (*r*_*g*_) between the common liability GWA (filtered according to Q_SNP_ *p*<5×10^−8^) and 41 other phenotypes, including 35 other traits (highlighted in blue) and the six individual substance use phenotypes used to derive the common liability (highlighted in grey). The asterisk indexes significant genetic correlations after correction for multiple testing (false discovery rate controlled at 5%, corrected for 41 tests). The full set of results is reported in **sTable 12** (Supplement).

### Evaluation of the causal relationships implied by a common liability model

**Figure 5** displays the results from Mendelian Randomization (MR) analyses, assessing paths running from the common liability to the individual substance use phenotypes. Using 42 Q_SNP_-filtered LD-independent SNPs from the common liability GWA, the MR findings provide support for a causal interpretation of the initial descriptive common liability model (cf. **Figure 1b**) – that is, the common liability increases the risk of addiction to nicotine, alcohol and cannabis. More specifically, the loadings obtained from the genomic factor model of the common heritable were recovered using genetic markers instrumenting the common liability. As shown, the standardized causal effects obtained in MR were comparable to the factor loadings of the indicators (highlighted in red in **Figure 5**), as evident for measures of dependence [mean MR estimate: 0.79 (0.10 SD)] and measures of frequency of substance use [mean MR estimate: 0.33 (0.12 SD)]. The hypothesized structural model also asserts absence of causal effects between indicators belonging to a different class of substance (e.g., cannabis dependence → alcohol dependence), which was in line with our MR results. Finally, reverse causation (effects of the specific substance use indicators on the common liability to addiction) was indicated for three of the indicators. Further discussion on the interpretation of reverse causation in this context is included in the Supplement, together with the full set of MR results (cf. **sTable 13**).

**Figure 5.**
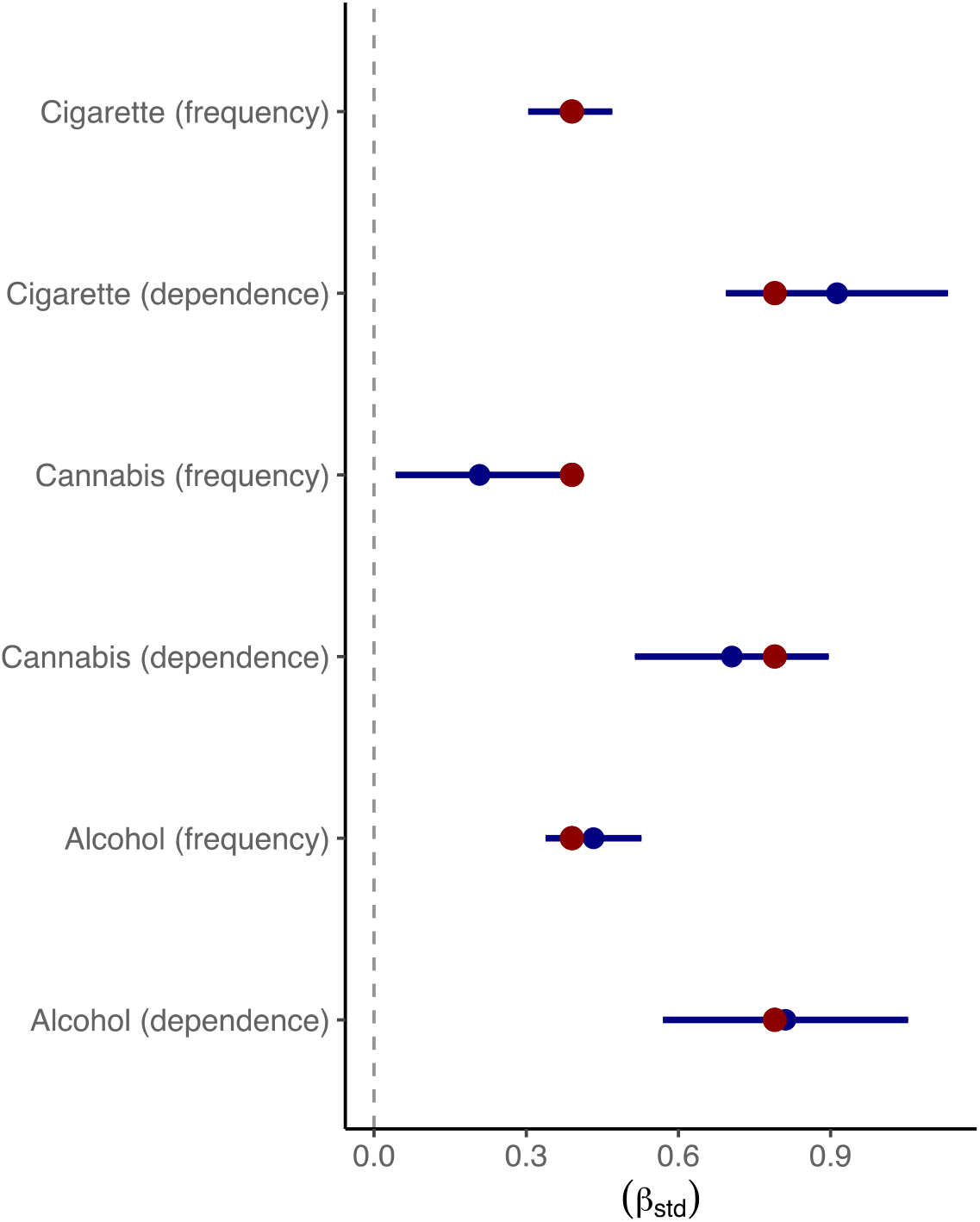
Mendelian Randomization analysis assessing causality between the common liability and the individual substance use phenotypes Shown are the standardized beta coefficients (β_std_) obtained from Mendelian Randomization (MR) analysis assessing the effects of the common liability on the six individual substance use phenotypes. Included were 42 genome-wide significant genetic variants (*p*<5×10^−8^) operating through the common liability (Q_SNP_ *p*>5×10^−8^) as instruments for the exposure. The red dots indicate the standardized loadings per substance use phenotype on the common liability as estimated in the structural model shown in **Figure 1B**. The full set of MR results can be found in **sTable 13**.

## Discussion

To dissect shared from non-shared genetic liability to addiction to nicotine, alcohol and cannabis, we conducted a multivariate genome-wide association (GWA) study of a common heritable liability to addiction. The top genetic variant operating through the common liability (rs10750025, located on the dopamine receptor D2 [*DRD2*] gene) provides support for the role dopamine in risk of addiction. Functional follow-up of common liability-associated genes further highlighted the role of widespread neuronal signalling pathways and neurotransmitter functioning beyond dopamine, such as GABAergic and glutamatergic pathways. Brain areas implicated in the common liability to addiction spanned limbic and cortical areas involved in reward, motivation, memory and cognitive control. The genetic overlap between the common liability and other complex traits was most prominent for other measures of addiction (e.g., cocaine and heroin use), as well as impulsive behaviours (e.g., risk-taking, ADHD) and mood (e.g., depression, insomnia). For cigarette and alcohol use, risk genes not operating via the common liability translated into specific pharmacogenomic pathways, such as nicotinic acetylcholine receptor functioning. Distinct pathways for cannabis use were, however, not identified.

### Shared and non-shared genetic risks involved in risk of addiction to nicotine, alcohol and cannabis

In line with existing evidence, we found substantial genetic correlations between measures of cigarette, alcohol and cannabis use. This allowed us to model the common heritable liability to addiction, which explained substantial variance (average=39%) in genetic liabilities to individual addiction phenotypes. *DRD2* was identified as the lead gene operating via the common liability – a pathway believed to be a common mechanism by which addictive substances exert their acute pleasurable effects. *DRD2* in particular is a frequently studied gene implicated in addictive behaviours, given its central role in modulating the dopamine reward system that mediates the reinforcing effects of addictive substances. Indeed, *DRD2* has been identified in numerous genome-wide studies on cigarette^7,8^, alcohol^7,8,20^ and cannabis use^8^. Other notable genes linked to the common liability included brain-derived neurotrophic factor (*BDNF*), corticotropin-releasing hormone receptor 1 (*CRHR1*), G protein-coupled receptor 1 (*GPCR1*), Ankyrin Repeat and Kinase Domain Containing 1 (*ANKK1*) or cAMP responsive element binding protein 3 like 1 (*CREB3L*). While these genes have already been scrutinized in studies using animal models and candidate gene approaches^64,65^ aiming to understanding the molecular basis of addiction, this is the first study confirming their involvement using a ‘hypothesis-free’ (i.e., genome-wide) approach.

Our results also highlight the role of neural signalling pathways involved in the common heritable liability to addiction, particularly synaptic functioning and a range of neurotransmitter systems beyond dopamine (GABA, glutamate, serotonin). Indeed, while dopaminergic mechanisms have been the traditional focus in addiction research, a growing body of research is now assessing the role of wider-ranging and interconnected neurotransmission systems in addiction vulnerability, involving GABAergic, glutamatergic and serotoninergic projections that contribute to modulating reward reinforcement and drug-seeking behaviour^66–68^. In line with this, enrichment analysis implicated the central nervous system and a network of brain areas in the common liability, including circuits involved in the processing of information related to reward (limbic structures), motivation (basal ganglia), memory (hippocampus) and cognitive control (frontal lobe areas). Since the discussed pathways were most prominently related to the common liability, rather than the individual substance use liabilities, the identified pathways may reflect common neural substrates characterizing addiction vulnerability. Indeed, genetic variants associated with the individual cigarette and alcohol use phenotypes showed a different molecular footprint when compared to variants associated with the common liability. Substance-specific genetic risk, defined as genetic risk not operating via the common liability, was most notably expressed in pharmacological pathways relevant to nicotine and alcohol. For cigarette use, six of the 16 central nervous system-expressed nicotinic acetylcholine receptor genes were associated with at least one of the cigarette use phenotypes. For alcohol use, four genes belonging to the alcohol dehydrogenase family associated with at least one of the alcohol use phenotypes. The existence of these substance-specific genetic risks may help explain why some individuals become addicted to either nicotine or alcohol. For cannabis use, there was less clear evidence for distinct genetic risk, as only one cannabis-associated gene (*GULOP*) did not operate through the common liability.

Finally, it is assumed that the brain reward pathways partly link to addictive behaviours via some intermediate complex behaviours, such as risk-taking, sensation seeking or impulsivity. While this remains to be formally tested, this idea corroborates with our findings of genetic correlations between the common liability and maladaptive behaviours, including ADHD, risk-taking and cocaine and heroin dependence. We also found large genetic correlations between the common liability with internalising symptoms, including depression and insomnia.

### Implications for the aetiology of addiction

Before jumping to aetiological conclusions regarding the shared and non-shared risks involved in addiction, it is important to assess if the core causal claims imposed by the common liability model hold true. Bi-directional MR was used to evaluate key assumptions, namely (1) the common liability to addiction has direct effects on all its indicators (i.e., the individual substance use phenotypes) and (2) there is no reverse causation. A third claim typically made by strict latent factor models asserts that there are no mutual direct effects between the individual indicators. In our study, this assumption was relaxed as we allowed for residual correlations between phenotypes belonging to the same class of substance (e.g., between frequency of cigarette use and cigarette dependence). Rather, our structural model posits that (3) all observed correlations between phenotypes belonging to a *different* class of substance are explained by the common liability. Overall, MR findings provided support for assumption (1), as causality ran from the common liability to all of the individual substance use phenotypes. While effects in the reverse direction were also present for three indicators – at odds with assumption (2) – this may reflect unaccounted pleiotropy (cf. Supplement for further discussion on this point). In line with assumption (3), there were no causal relationships between phenotypes belonging to a different class of substance. Together, since core assumptions are met (common liability → individual phenotypes), the results suggest that a common liability to addiction may usefully capture most relationships between substances. Embracing such conceptualization would have important implications for intervention. First, targeting modifiable features of the common liability should reduce risk of addiction to nicotine, alcohol and cannabis. For example, pharmacological treatments targeting dopamine, glutamate and GABA function may reduce craving and the euphoric/rewarding responses to cigarettes, alcohol and cannabis^69–74^. Second, interventional targeting of only one specific class of substance (e.g., nicotine) unlikely leads to reductions in use of another class (e.g., alcohol). This conclusion is somewhat inconsistent with previous evidence in rodents showing reductions in alcohol use following the administration nicotinic treatments (e.g. varenicline^75,76^), although evidence from RCTs in humans is mixed^77,78^ and efficacy may not translate into the long-term^79^.

### Future directions

An important goal for future work would be to extend this multivariate analysis to a broader array of substance classes (e.g., cocaine, opiate) and addiction phenotypes (e.g., tolerance, craving, withdrawal, relapse), once the required GWA data becomes available. Such efforts will allow exploration of more fine-grained structural models. Furthermore, multivariate approaches as employed here are not just important in terms of GWA discovery, but also essential to reducing biases; in our study, a substantial proportion of GWA-significant SNPs associated with the individual substance use phenotypes appeared to be mediated by the common heritable liability. As such, future GWA studies powerful enough to detect small genetic effects will likely tag an increasing number of SNPs with horizontal pleiotropic effects when examining addiction phenotypes (i.e., direct effects on several phenotypes). As such, modelling heritable latent factors as done in this study, and/or accounting for its contribution as recently proposed^80,81^ is therefore paramount when using genetically informed causal inference methods that are sensitive to the presence of heritable confounding.

Taken together, our results confirm that a common heritable liability partially explains the high co-occurrence of addiction to nicotine, alcohol, and cannabis. Functions of the implicated genes converged on broad central nervous system pathways beyond the dopaminergic pathways long-hypothesised in risk of addiction.

## Data Availability

Summary statistics of the common liability GWA analysis will be available upon publication of this work. References to all publicly available summary statistic files included in this work are listed in sTable 1.

https://github.com/TabeaSchoeler/TS2021_CommonLiabAddiction

## Data access

Summary statistics of the common liability GWA analysis will be available upon publication of this work. References to all publicly available summary statistic files included in this work are listed in **sTable 1**.

## Code availability

The code used to conduct the analyses presented in this work is available on GitHub (https://github.com/TabeaSchoeler/TS2021_CommonLiabAddiction)

## URLs

MsigDB data (https://www.gsea-msigdb.org/gsea/index.jsp)

PASCAL (https://www2.unil.ch/cbg/index.php?title=Pascal)

G:Profiler (https://biit.cs.ut.ee/gprofiler/page/r/)

PhenoScanner (https://github.com/phenoscanner/phenoscanner)

TwoSampleMR (https://mrcieu.github.io/TwoSampleMR/)

GenomicSEM (https://github.com/MichelNivard/GenomicSEM)

DEPICT (https://data.broadinstitute.org/mpg/depict/)

Neale Lab UKBB summary statistics (http://www.nealelab.is/uk-biobank/)

GWAS ATLAS (https://atlas.ctglab.nl/)

## Acknowledgements

This study would not have possible without the use of publicly available genome-wide summary data and software tools. The authors gratefully acknowledge these resources, and thank the research participants, the research teams and institutions that have contributed to this research. We are grateful to Dr. Judit Cabana-Domínguez and Prof. Bru Cormand for providing the results from the cocaine dependence genome-wide association study.

