## Supplementary material for "Novel insights into the common heritable liability to addiction: a multivariate genome-wide association study"

Table of content

SMETHODS 2

**Estimation of the effective sample size of the common liability genome-wide association analysis** **2**

**Enrichment analysis** **2**

**Standardization of beta estimates** **2**

SFIGURES 3

**sFigure 1.** Manhattan and QQ-plot of the uni- and multivariate genome-wide association analyses 3

**sFigure 2.** PASCAL pathway enrichment analysis of genes associated with the common heritable liability 5

**sFigure 3.** Bi-directional Mendelian Randomization analysis assessing causality between the common liability and the substance use phenotypes 6

SREFERENCES 6

#### sMethods

##### Estimation of the effective sample size of the common liability genome-wide association analysis

To derive an estimate of the effective sample size for the overall genome-wide association (GWA) analysis, we followed the formula proposed by Mallard et al.[1]:

$$n_j = \frac{(Z_j/\beta_j)^2}{2 \times MAF_j\,(1 - MAF_j)}$$

$$N_{eff} \approx \frac{1}{m} \sum_{MAF=a}^{b} n_j$$

Where $n_j$ is the unknown effective sample size per SNP $j$, $Z_j$ is the Z statistic of $SNP_j$, $MAF_j$ is the minor allele frequency $SNP_j$ and $\beta_j$ is the effect estimate obtained from the GWA on the common liability for $SNP_j$. As this formula may produce incorrect estimates for SNPs with low MAF, the effective sample size (Neff) was estimated as an average of $n_j$ based on $m$ SNPs with MAF $a$ > 10% and MAF $b$ < 40%.

##### Enrichment analysis

We used Data-driven Expression-Prioritized Integration for Complex Traits (DEPICT[2]) to test for tissue/cell type enrichment. DEPICT relies on gene expression data derived from 77,840 samples (gene expression microarrays) and public pathway annotation datasets (e.g., GO, KEGG, REACTOME, MGI). DEPICT was applied to a set of a set of LD-independent SNPs ($r^2$<0.05 within 500 kb) outside genome-wide significance ($p$<5×10$^{-5}$), using the default settings. The common liability GWA results were filtered according to $Q_{SNP}$ prior to its analysis using DEPICT, including only SNPs that did not show heterogenous effects ($Q_{SNP}$ $p \geq 5\times10^{-8}$).

Pathway SCoring ALgorithm (PASCAL)[3] was used to test for enrichment of all SNPs, using three gene sets (BIOCARTA, KEGG, REACTOME) curated by the Molecular Signatures Database (MSigDB[4]) and gene sets defined by DEPICT. Results obtained from PASCAL were corrected for multiple testing using false discovery rate (FDR) correction. FDR correction was applied separately to two sets of results, namely the $\chi^2$ $p$-values obtained from enrichment analysis using MSigDB[4] (1077 sets) and enrichment analysis using gene sets defined by DEPICT (14462 sets). We used the default settings set by PASCAL, using the sum option (based on the average association signal across a region) to run conduct gene scoring. The common liability GWA was filtered according to $Q_{SNP}$ prior to its analysis using DEPICT, including only SNPs that did not show heterogenous effects ($Q_{SNP}$ $p \geq 5\times10^{-8}$).

##### Standardization of beta estimates

To facilitate comparability of the Mendelian Randomization results, the beta estimates and corresponding standard errors of all included SNPs were standardized based on the following formula:

$$Z_j = \frac{\beta(SNP_j)}{SE(SNP_j)}$$

$$\beta_{STD}(SNP_j) = \frac{Z_j}{\sqrt{N_j}}$$

$$SE_{STD}(SNP_j) = \sqrt{\frac{1}{N_j}}$$

Where $Z_j$ is the Z statistic of $SNP_j$, $N_j$ is the GWA sample size per $SNP_j$, β (SE) is the unstandardized effect estimate (corresponding standard error) of $SNP_j$ with the phenotype and $\beta_{STD}$ ($SE_{STD}$) is the standardized effect (corresponding standard error) of $SNP_j$ with the phenotype. For binary traits, $N_j$ indexes the effective sample size of the GWA per $SNP_j$, estimated as:

$$Neff = \frac{2}{\frac{1}{n_{case}} + \frac{1}{n_{control}}}$$

### sFigures

sFigure 1. Manhattan and QQ-plot of the uni- and multivariate genome-wide association analyses

**Column 1.** Manhattan plots of the SNP effects obtained from the multivariate genome-wide association analysis on the common heritable liability, as well as the SNP effects from the univariate genome-wide association analyses on the individual substance use phenotypes. Labels are provided for the LD-independent genome-wide significant SNPs (i.e., SNPs above the horizontal line, with $p<5\times10^{-8}$) and gene names obtained through positional mapping. The x-axis refers to chromosomal position, the y-axis refers to the *p*-value on a -log10 scale. Genetic variants coloured in red index variants that showed heterogeneous effects across the individual cigarette, alcohol and cannabis use phenotypes ($Q_{snp}$ $p<5\times10^{-8}$), indicating that their effects operate not entirely through the common liability. Genetic variants coloured in blue index variants that did not show heterogeneous effects across the individual cigarette, alcohol and cannabis use phenotypes ($Q_{snp}$ $p\geq5\times10^{-8}$), indicating that their effects are likely to operate through the common liability.

**Column 2.** QQ-plot of the observed and expected p-values for each of the genome-wide association results.

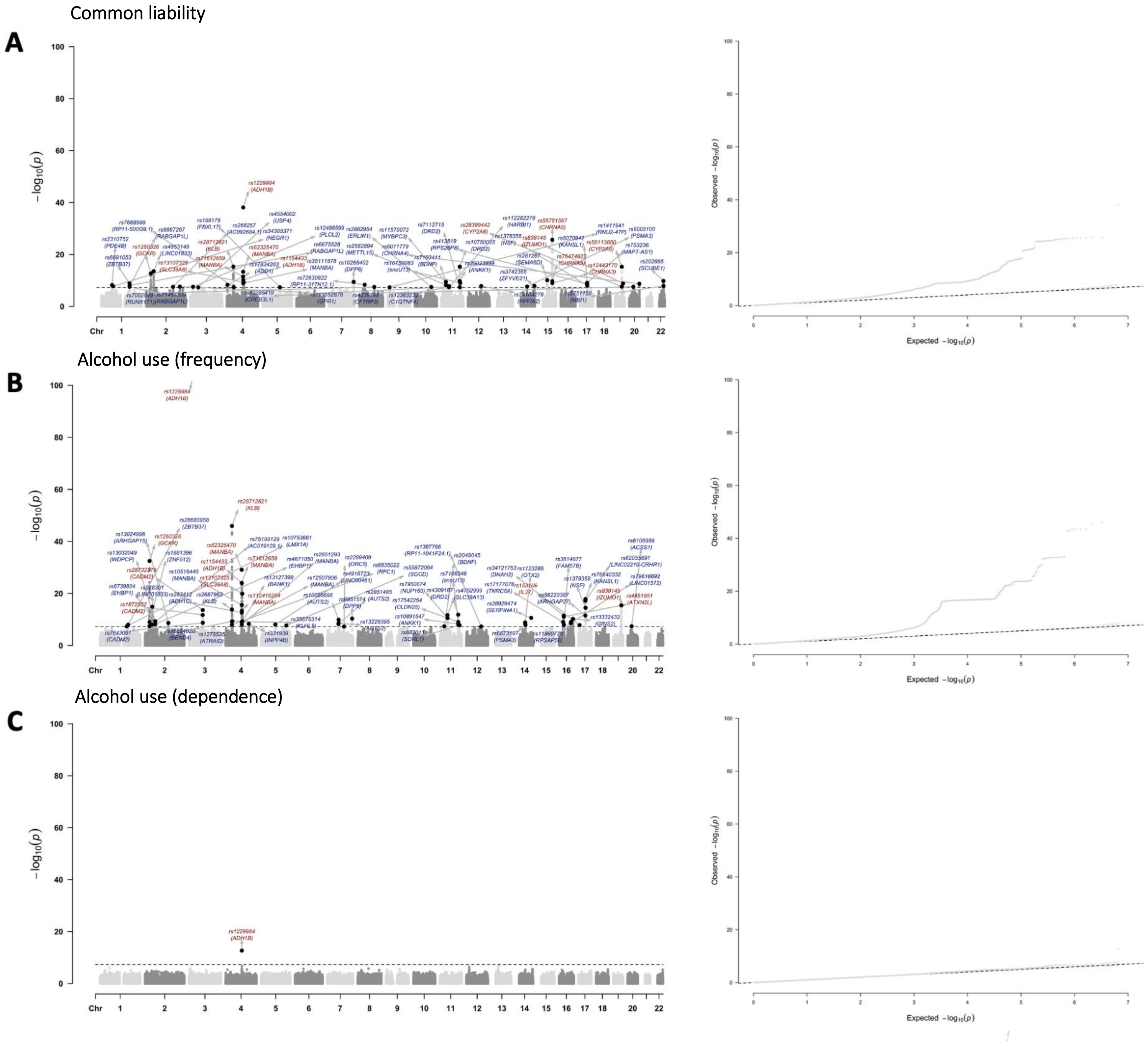

**D** Cigarette use (frequency)

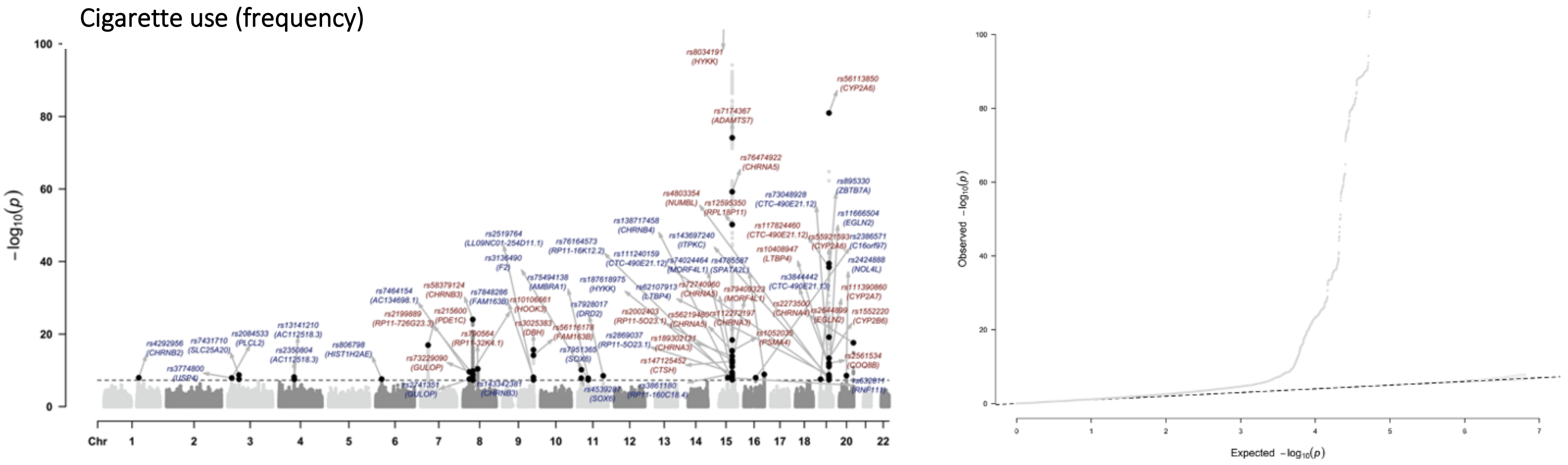


**E** Cigarette use (dependence)

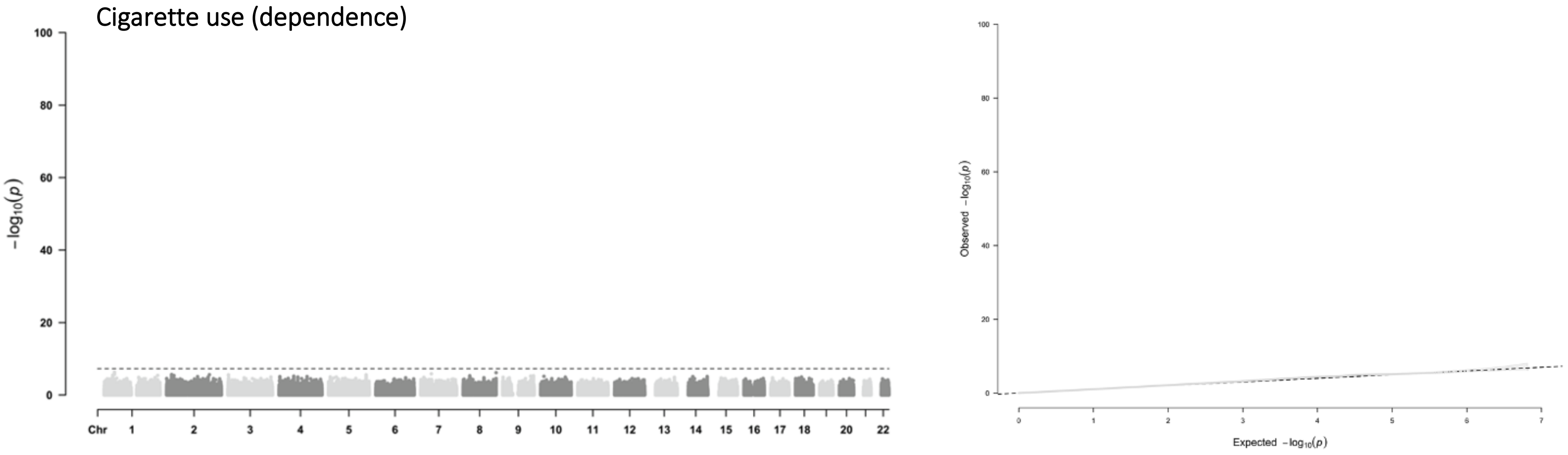


**F** Cannabis use (frequency)

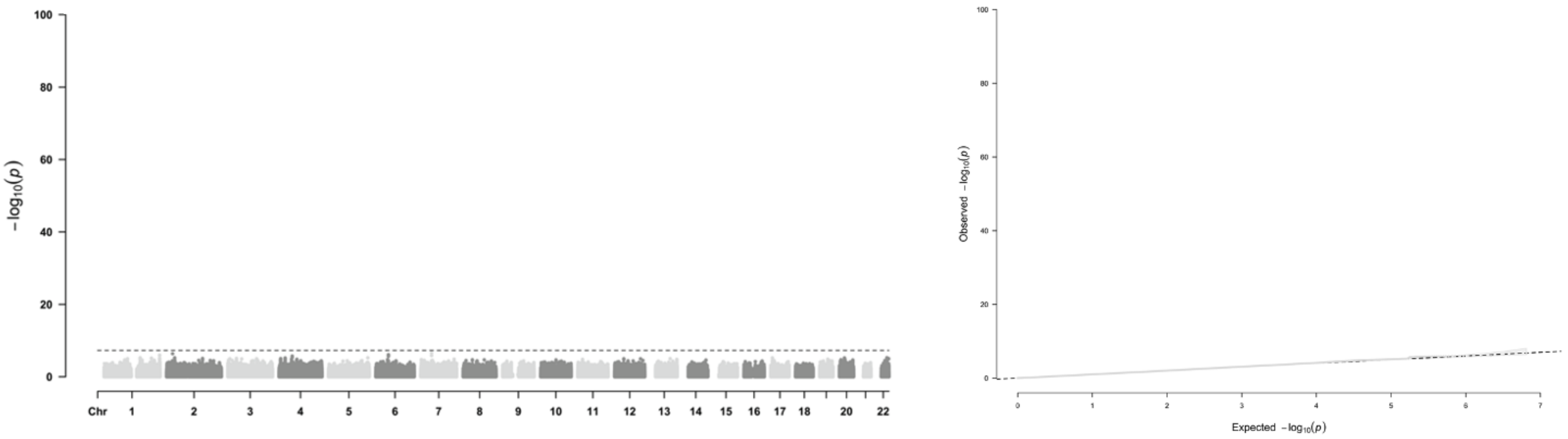


**G** Cannabis use (dependence)

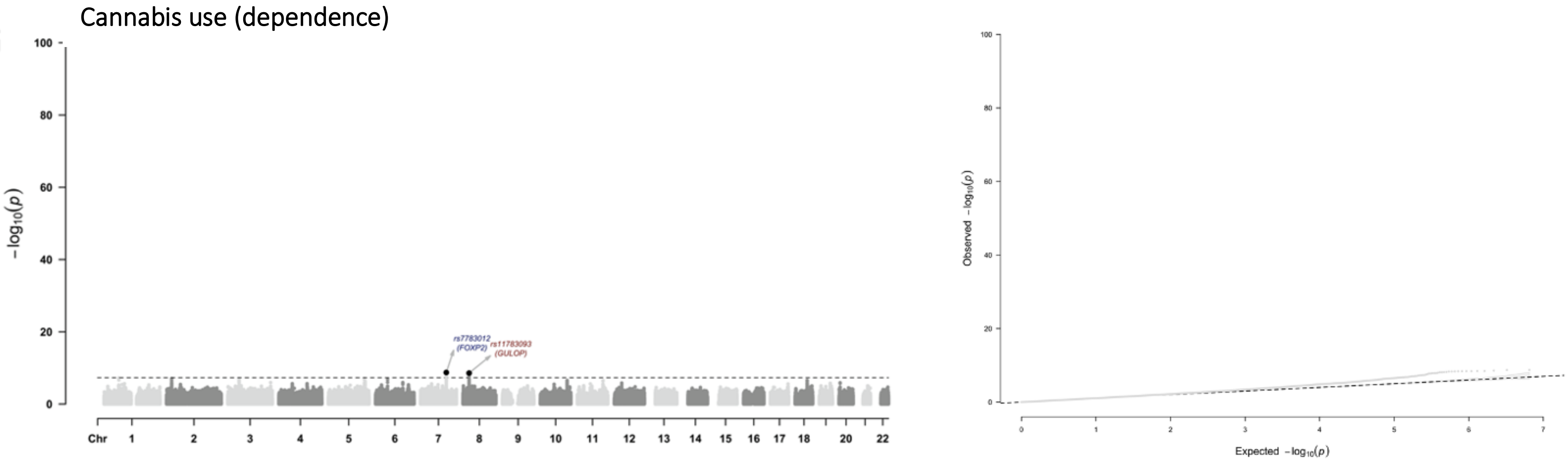

*Evaluation of the causal relationships implied by the common liability theory*

**sFigure 3.** displays the results from Mendelian Randomization (MR) analyses, assessing paths from the common liability to the individual substance use phenotypes (direct causation) and the reverse (reverse causation), from the indicator to the common liability. Using 42 $Q_{SNP}$-filtered LD-independent SNPs from the common liability GWA, the MR findings provide validation for the factor loadings of the initial common liability model (cf. Figure 1b, main manuscript) – that is, the loadings obtained from genome-wide analyses were reproduced using a number of genetic instruments indexing the common liability. Reverse causation, was most strongly implicated for two of the indicators (frequency of cigarette and alcohol use) and, albeit to a lesser extent, alcohol dependence ($p_{IVW}$=0.044). This result reflects either 1) true causal effects (which would violate the model assumptions) or 2) biased effects resulting from the inclusion of invalid instruments. With respect to 1), causal effects of the indicators may be interpreted as pathways where the use of specific drugs (e.g., cigarettes, alcohol) would sensitize common pathways (e.g., dopamine), which in turn would increase the risk of using other drugs. With respect to 2), some of the selected instruments for measures of frequency of use may affect the common liability through pathways other than the exposure they are indexing, which would violate the MR assumptions and result in biased MR estimates. Since the common liability explains less variance for measures of frequency of use compared to measures of dependency, larger residual substance-specific effects exist for measures of frequency of use. As such, reciprocal causal pathways may simply reflect the resulting pleiotropy, where substance-specific effects correlate with variant effects estimated for the common liability.

**sFigure 3.** Bi-directional Mendelian Randomization analysis assessing causality between the common liability and the substance use phenotypes

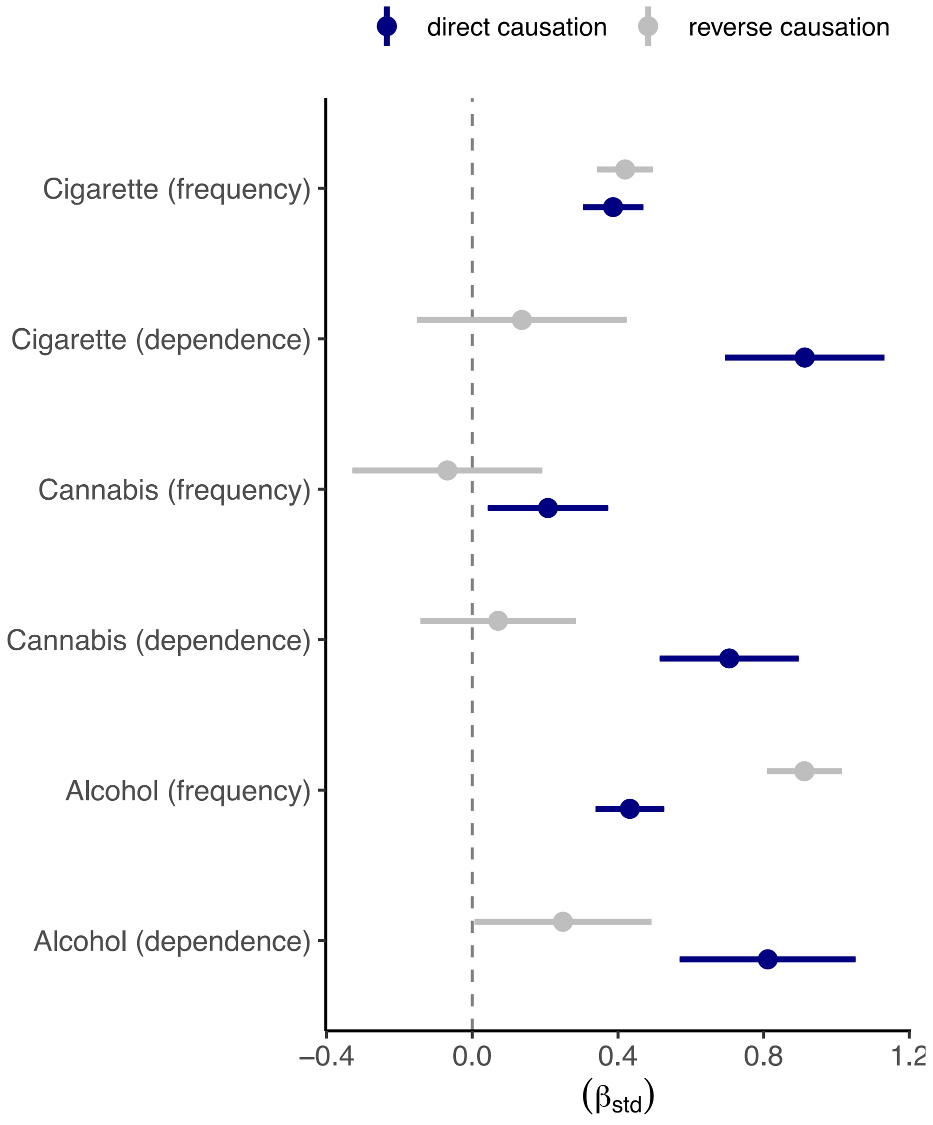


Shown are the standardized beta coefficients obtained from Mendelian Randomization (MR) analysis assessing the bi-directional relations between the common heritable liability to addiction and the six individual substance use phenotypes. Direct causation was estimated as the effects of the common liability on the substance use phenotypes, including n=42 genome-wide significant genetic variants ($p<5\times10^{-8}$) operating through the common liability ($Q_{snp}$ $p$-$\geq 5\times10^{-8}$) as instruments. Reverse causation was assessed by estimating the effects of the individual substance use phenotypes on the common liability, including only genome-wide significant genetic variants that did not operate through the common liability ($Q_{snp}$ $p<5\times10^{-8}$) as instruments. In instances where the GWA data did not contain genetic variants reaching genome-wide significance, we selected the top 10 LD-independent SNPs as instruments. A description of the instruments used in the analysis, as well as full set of MR results can be found in **sTable 13**.

#### sReferences


1. Mallard TT, Linnér RK, Okbay A, et al. Not just one p: Multivariate GWAS of psychiatric disorders and their cardinal symptoms reveal two dimensions of cross-cutting genetic liabilities. *bioRxiv*. Published online January 1, 2019:603134. doi:10.1101/603134
2. Pers TH, Karjalainen JM, Chan Y, et al. Biological interpretation of genome-wide association studies using predicted gene functions. *Nat Commun*. 2015;6(1):5890. doi:10.1038/ncomms6890
3. Lamparter D, Marbach D, Rueedi R, Kutalik Z, Bergmann S. Fast and Rigorous Computation of Gene and Pathway Scores from SNP-Based Summary Statistics. Listgarten J, ed. *PLOS Comput Biol*. 2016;12(1):e1004714. doi:10.1371/journal.pcbi.1004714
4. Subramanian A, Tamayo P, Mootha VK, et al. Gene set enrichment analysis: A knowledge-based approach for interpreting genome-wide expression profiles. *Proc Natl Acad Sci*. 2005;102(43):15545-15550. doi:10.1073/pnas.0506580102
